# Mapping genetic determinants of 184 circulating proteins in 26,494 individuals to connect proteins and diseases

**DOI:** 10.1101/2021.08.03.21261494

**Authors:** Erin Macdonald-Dunlop, Lucija Klarić, Lasse Folkersen, Paul R.H.J. Timmers, Stefan Gustafsson, Jing Hua Zhao, Niclas Eriksson, Anne Richmond, Stefan Enroth, Niklas Mattsson-Carlgren, Daria V. Zhernakova, Anette Kalnapenkis, Martin Magnusson, Eleanor Wheeler, Shih-Jen Hwang, Yan Chen, Andrew P Morris, Bram Prins, Urmo Võsa, Nicholas J. Wareham, John Danesh, Johan Sundstrom, Bruna Gigante, Damiano Baldassarre, Rona J. Strawbridge, Harry Campbell, Ulf Gyllensten, Chen Yao, Daniela Zanetti, Themistocles L. Assimes, Per Eriksson, Daniel Levy, Claudia Langenberg, J. Gustav Smith, Tõnu Esko, Jingyuan Fu, Oskar Hansson, Åsa Johansson, Caroline Hayward, Lars Wallentin, Agneta Siegbahn, Lars Lind, Adam S. Butterworth, Karl Michaëlsson, James E. Peters, Anders Mälarstig, Peter K. Joshi, James F. Wilson

## Abstract

We performed the largest genome-wide meta-analysis (GWAMA) (Max N=26,494) of the levels of 184 cardiovascular-related plasma protein levels to date and reported 592 independent loci (pQTL) associated with the level of at least one protein (1308 significant associations, median 6 per protein). We estimated that only between 8-37% of testable pQTL overlap with established expression quantitative trait loci (eQTL) using multiple methods, while 132 out of 1064 lead variants show evidence for transcription factor binding, and found that 75% of our pQTL are known DNA methylation QTL. We highlight the variation in genetic architecture between proteins and that proteins share genetic architecture with cardiometabolic complex traits. Using *cis*-instrument Mendelian randomisation (MR), we infer causal relationships for 11 proteins, recapitulating the previously reported relationship between PCSK9 and LDL cholesterol, replicating previous pQTL MR findings and discovering 16 causal relationships between protein levels and disease. Our MR results highlight IL2-RA as a candidate for drug repurposing for Crohn’s Disease as well as 2 novel therapeutic targets: IL-27 (Crohn’s disease) and TNFRSF14 (Inflammatory bowel disease, Multiple sclerosis and Ulcerative colitis). We have demonstrated the discoveries possible using our pQTL and highlight the potential of this work as a resource for genetic epidemiology.

## Introduction

Proteins are the key functional elements in the body and are instrumental in most biological processes including, growth, repair, transport and signalling. Dysregulation of proteins circulating in the blood is often observed in disease and, moreover, is often part of the causal pathway, making them ideal candidate drug targets. The plasma proteome, which consists of both proteins which are actively secreted and passively leaked from cells, is an attractive and accessible system to study. As samples are easy to store, collection is minimally invasive for study participants, and hundreds to thousands of molecules can be measured, plasma proteins have been investigated as biomarkers for numerous diseases^1^. The recent advances in targeted proteomics technologies have allowed thousands of circulating plasma protein levels to be measured simultaneously, even in large sample sizes^2–9^. Uncovering relationships between protein biomarkers and disease has the potential to aid in prediction of risk, diagnosis and development of new therapies for disease^10^. Cardiovascular disease (CVD)-related proteins are of particular interest as CVD was the leading cause of morbidity and mortality globally in 2019^11^, being responsible for 18.6 million deaths and 393 million disability adjusted life years (DALYs).

As circulating plasma protein levels are partly heritable^12^, genome-wide association studies (GWAS) have been used to discover genetic loci that are associated with regulation of protein levels; protein quantitative trait loci (pQTL)^3,4,13–17^. Previous pQTL studies have uncovered potential mechanisms of action for how common genetic variants affect circulating protein levels^4,18^.

Using Mendelian Randomisation (MR) to assess potential causal relationships between biomarkers and disease phenotypes has become an increasingly utilised approach for drug target discovery and validation and has also successfully predicted outcomes of randomised controlled trials (RCTs)^19^. Despite many associations between levels of circulating biomarkers and various diseases in the literature, positing causal roles for these biomarkers has only been possible through the application of methods such as MR. The study of the genetics of circulating biomarkers such as plasma protein levels therefore has the capacity to uncover pathways, disease aetiology, therapeutic targets and biomarkers to aid detection and diagnosis of disease^10^. Unlike GWAS of complex traits, targets highlighted studying the plasma proteome are themselves directly actionable.

Previous large GWAS of plasma protein levels have discovered hundreds of associated loci, uncovered mechanisms for pQTL, causal relationships between proteins and diseases and posited how plasma protein levels may act to influence disease risk^3,4,7,8,17,18,20^. In order to maximise the potential for pQTL discovery and MR to find causal associations with disease and build on previous work, we performed genome-wide meta-analysis with the largest sample size for 184 cardiovascular-related plasma proteins. We uncovered insights into the genetic architecture of these proteins, indicating potential mechanisms for pQTL from altering gene expression and beyond. Using a broad exploratory analysis, we demonstrate the power of pQTL as genetic instruments in MR and highlight potential causal relationships between proteins and disease. These results suggest putative drug targets and repurposing opportunities. With this work we have created a resource of pQTL data that will aid the field of genetic epidemiology and provide tools for targeted experiments in the future.

## Results

### Discovery of Protein Quantitative Trait Loci

We performed genome-wide meta-analyses (GWAMA) of the levels of 184 cardiovascular-related plasma protein levels measured by the Olink proximity extension assay in a maximum of 26,494 individuals of European ancestry from 18 cohorts. We identified 1,073 significant protein-locus associations (*cis*: p<1.18 × 10^−7^, *trans*: p<5.55 × 10^−10^, where *cis* was defined as ±1 Mb flanking the protein-coding region). After performing conditional analysis, we report a further 235 conditionally-independent protein-variant associations. In total we found 1,308 significant lead variant-protein associations, 288 *cis*-associations and 1,020 in *trans* (Figure 1a, Supplementary Table 1). This equates to the discovery of 592 independent loci significantly associated with the levels of at least one protein (Supplementary Table 2).

**Figure 1.**
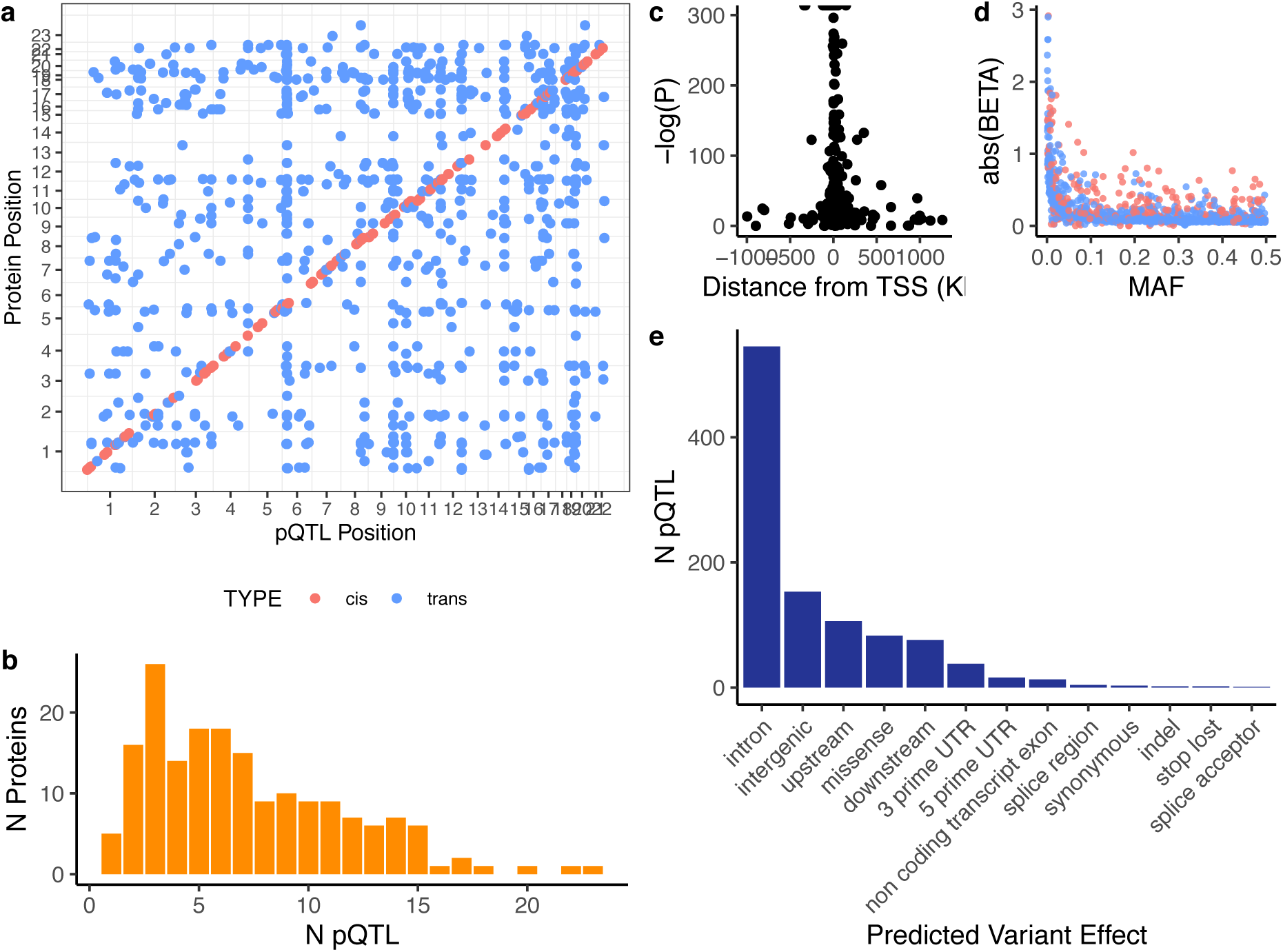
Significantly associated loci from GWAMA of 184 proteins. a) Points indicate the genomic position of 1,308 significant pQTL against the genomic position of the transcriptional start site (TSS) of the gene encoding the protein that the variant is associated with. Colour indicates if the variant is a cis- or trans-pQTL. Cis-is defined as any variant within ±1 Mb of the coding region of the gene encoding the protein, trans-defined as any variant outside that region. b) Histogram of the counts of significant pQTL per protein. c) Relationship between -log P-value and distance of each cis-pQTL from the TSS of the gene. d) Magnitude of effect size (absolute beta) shows a typical L-shaped relationship with minor allele frequency (MAF) of our pQTL (cis in blue, trans in pink). e) The frequency of predicted effect of the sentinel variants.

Only two proteins, growth hormone 1 (GH) and Inhibitor of nuclear factor kappa B kinase regulatory subunit (NEMO), had no significant pQTL. For an additional 16 proteins we found no significant *cis*-signals (ACE2, CCL22, CD40-L, CD93, Ep-CAM, GDF-2, HAOX1, ICAM-2, IL-6, ITGB1BP2, MB, PDGF subunit B, PECAM-1, SRC, t-PA, VEGF-D). For five of the above eighteen proteins, ACE2, CD40-L, NEMO, ITGB1BP2 and VEGF-D, this is expected as they are encoded on the X chromosome, which was not studied here. For the remaining 13 proteins, the minimum p-value and *cis* regions are shown in Supplementary Figure 1. Altogether, we report significant *cis*-pQTL for 92.7% of the plasma proteins (where *cis*-regions were tested). 182 out of the 184 proteins analysed had a pQTL, 155 proteins had both *cis*- and *trans*-pQTL, 11 *cis*-only and 16 *trans*-only.

The majority of proteins had 6 or fewer significant pQTL (Median N pQTL = 6), with 3 proteins (CD163, CTSL1 and RAGE) having more than 20 (Figure 1b). In general proteins with multiple significant pQTL had 1-3 *cis*-associated pQTL with any additional associated loci being in *trans* (Supplementary Figure 2). Interestingly, 241 loci contained pQTL for more than one protein, with the *HLA* and *ABO* regions being associated with the most proteins (Supplementary Table 3). We saw a distinct pattern with our significant *cis*-associated variants such that variants nearest the transcriptional start site (TSS) of the relevant gene displayed the strongest associations (Figure 1c). As seen for most complex traits, the magnitude of effect size increased with decreasing minor allele frequency (Figure 1d). Using single variant annotation from Ensembl variant effect predictor^21^ we found that 70% of our lead variants are either intronic or intergenic (Figure 1e).

Six hundred and twenty-one (47.5%) of our significant lead variants (or variants in LD, r^2^>0.5, with our lead variants) have previously been reported as genome-wide significantly associated with the relevant protein in previous GWAS of plasma protein levels (Details of previous studies summarised in Supplementary Table 4). Thus, 687 (52.5%) of our significant protein-variant associations are novel. We also reported 20 novel proteins with significant pQTL.

### Genetic Architecture of Plasma Protein Levels

Unlike traditional complex polygenic traits, many of the proteins have an extremely strong *cis*-association signal and have individual variants that explain relatively large proportions of variance in the phenotype. Having very few (or one) strong signals is rare outside of molecular phenotypes, as many weak signals with small effects are common in complex traits. We found that 75 of our 1,308 lead variants have estimated phenotypic variance explained (R^2^) of >5% (51 *cis* and 24 *trans*), with the highest being rs12141375:A, estimated to explain 32.7% of the variance in plasma CHIT1 levels (Supplementary Figure 3).

Standard methods for estimating single nucleotide polymorphism (SNP) heritability from association summary statistics assume a polygenic model that is unlikely to hold for proteins. We therefore calculated the heritability contributed by significant pQTL (pQTL component) and the remaining genome-wide SNP heritability (polygenic component), separately for each protein (Figure 2). The pQTL component was calculated as the sum of the estimated variance in protein level explained by the lead SNPs and the polygenic component estimated using LD-score regression^22,23^ (see methods for details). Estimates of total genetic component ranged from 2.9% for NEMO protein levels to 40.2% for CHIT1. Genetic architecture, however, varied across proteins with IL-6RA and CHIT1 protein levels having identified pQTL accounting for 96.5% and 93% of their SNP heritabilities, respectively. Conversely, the genetic components of NEMO and GH protein levels appear entirely polygenic, having no significant pQTL in this analysis.

**Figure 2.**
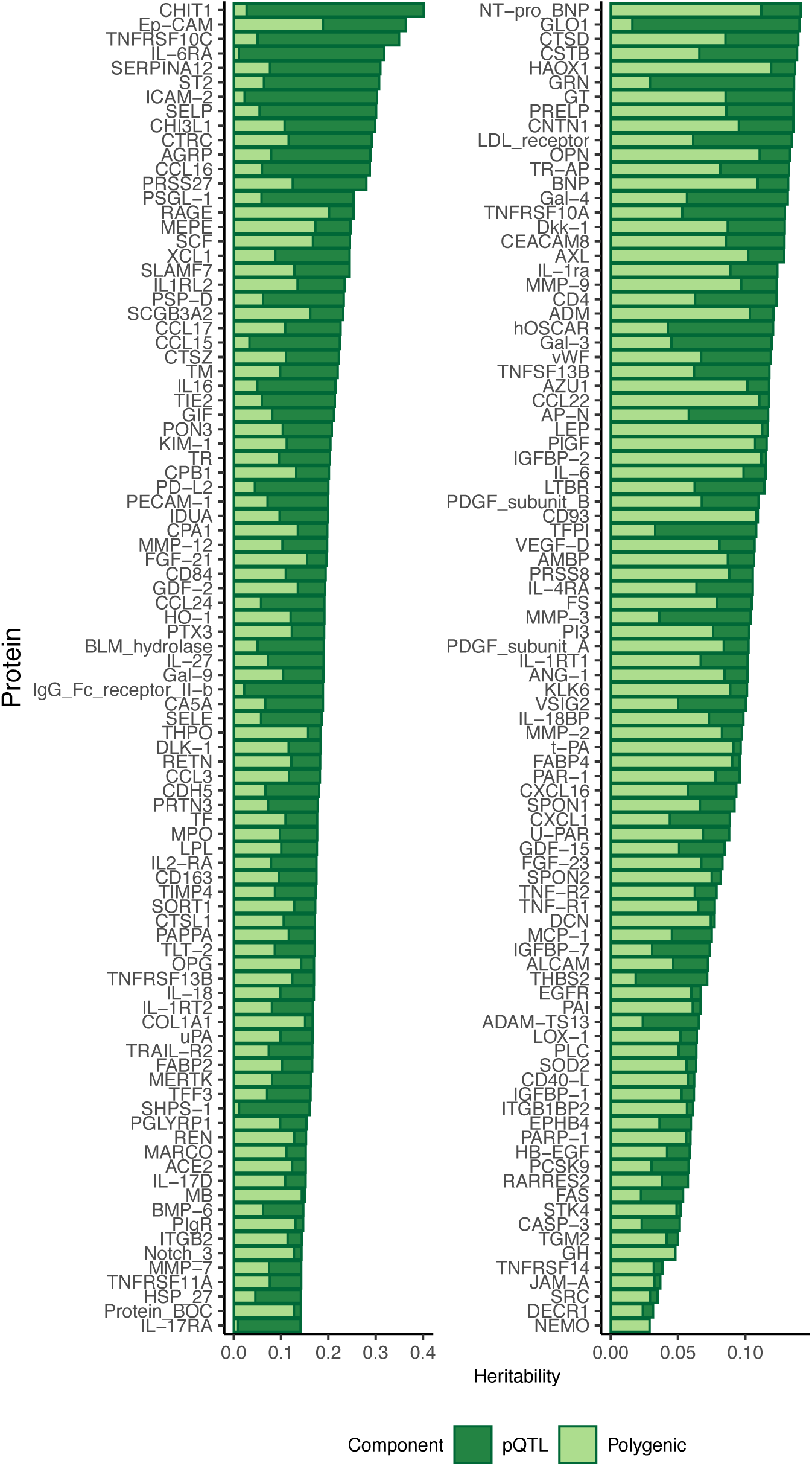
pQTL vs Polygenic Contribution to SNP heritability. The SNP heritability estimated for each protein, stratified by contributions from significant pQTL and polygenic effects. Polygenic: LDSC-estimated SNP heritability excluding variants indexed by the lead variants, pQTL: sum of the estimated variance in protein level explained by the lead variants (See Methods for details).

We observed that there is a relationship between the number of significant pQTL we found and the estimated SNP heritability, with increasing heritability estimates with increasing number of pQTL (Supplementary Figure 4).

### Colocalisation of pQTL & eQTL

We sought to uncover potential mechanisms by which our pQTL might act to influence the level of proteins circulating in plasma. Biologically, the most direct route biologically, would be for the significantly associated variants to affect protein levels by altering gene expression. 36.5% of the lead *cis*-variants have been previously reported as *cis*-expression QTL (eQTL) for the gene encoding the protein of interest (eQTLGen^24^, at 5% FDR (permutation-based)). However, for each of our pQTL, the lead variant (strongest association based on p-value) is not necessarily the causal variant. The lead variant commonly tags the signal for multiple variants in high LD, any of which could be the true causal variant.

To further define whether the signals were shared we used two different approaches. We first looked for evidence of gene expression mediating the effect of our pQTL on plasma protein abundances using summary based Mendelian Randomisation (SMR) and tested that these estimates were not due to linkage using the heterogeneity in dependent instruments (HEIDI) test^25^. We found associations between 1,371 transcripts and 168 proteins (PSMR<1.68 × 10^−7^, PHEIDI≥0.01) in at least one of four eQTL datasets (eQTLGen^24^,GTEx v7, Westra *et al*.^26^ and Cage^27^). The number of significant transcript-protein associations across different eQTL resources are shown in Supplementary Table 5, as well as how many of our proteins are associated with the expression level of the transcript encoding the protein, compared to other transcripts.

Secondly, to formally test whether the association signals with gene expression and protein level share the same causal variant or are driven by different variants, we looked for evidence of colocalisation. The Bayesian framework implemented by coloc^28^ assesses several hypotheses simultaneously by estimating separate posterior probabilities (PP) of the eQTL and pQTL i) sharing a single causal variant or ii) being caused by two independent variants. We found that 18 out of 220 testable *cis*-pQTL showed strong evidence of colocalisation (PP>0.8) with the *cis*-eQTL in whole blood using the eQTLgen summary statistics, with an additional 2 being likely to share a causal variant (PP>0.5). Using eQTL data for 48 different tissues from GTEx v7, we found that 40 out of 277 testable *cis*-pQTL colocalise (PP>0.8) with the *cis*-eQTL in at least one tissue, with 12 more being likely to share a causal variant (PP>0.5). The majority of pQTL which colocalise with eQTL do so in <6 tissues, however there are several that colocalise with eQTL across >20 tissues (Supplementary Figure 5). Interestingly, there are very few that appear to be tissue specific.

Both of these approaches share the caveat that they are unable to distinguish causality from pleiotropy. However, given that we are assessing the effect of genetic variants on gene expression and protein levels, the central dogma suggests these relationships are likely to be causal, but a definitive statement of causality for individual associations cannot be made using current methods.

### Other Potential Mechanisms

As not all pQTL appear to act by altering gene expression, we looked for other potential mechanisms of action. For some proteins we found *trans*-pQTL that map to that protein’s receptor or vice versa. For example, despite having no significant *cis*-pQTL, IL-6 has an extremely strong association in the IL-6 receptor (IL6-RA) region.

Given that 70% of our lead variants are intronic or intergenic, we next looked for existing annotation of regulatory function using RegulomeDB^29^ (Supplementary Figure 6a). Of the 1064 of our 1093 lead variants that have an entry in RegulomeDB, 50 (8 *cis* and 42 *trans*) were placed in category 1, meaning they are known eQTL with varying additional levels of support (e.g. transcription factor (TF) binding, TF motif, DNase footprint), for the variant being located in a functional region. 82 of our lead variants (25 *cis*, 56 *trans* & 1 both *cis* and *trans*, but for different proteins) were scored in category 2, meaning that despite not being known eQTL, variants have direct evidence of binding from ChIP-seq and DNase footprinting. These results suggest that a substantial minority of pQTL that are not yet reported as being significant eQTL influence TF-binding.

To uncover potential mechanisms for our *trans*-pQTL, we used annotation databases to see if *trans*-genes share pathways or are known to interact with the protein of interest. We defined *trans*-genes as all genes whose coding regions overlapped with a 1 Mb window centred on the lead variant of the *trans*-pQTL. We found that 85 of our *trans*-pQTL have a *trans*-gene with a known interaction with the protein of interest using STRINGdb^30^. Similarly, 37 *trans*-pQTL have a *trans*-gene that shares a common KEGG^31^ pathway with the gene encoding the protein of interest, 158 share common gene ontology (GO) terms and 816 have a *trans*-gene that is mentioned in a publication together with the protein of interest (Supplementary Table 6).

### pQTL Associated with Metabolites, DNA methylation levels & Complex Traits

Aside from affecting gene expression and plasma protein levels, our pQTL have also been previously associated with the levels of metabolites circulating in the plasma, with methylation of CpG dinucleotides and with complex traits. Using Phenoscanner^32^ we established that, of our 1093 unique lead variants: 96 have been reported as significantly (p<5 × 10^−8^) associated with circulating metabolite levels, 816 with DNA methylation and 547 with complex traits in GWAS (Supplementary Figure 6b).

The 547 lead variants reported in previous GWA studies, have been significantly associated with a broad range of phenotypes, from cardiovascular-related phenotypes to immune and inflammatory diseases (Supplementary Table 7). Lead variants were also associated with anthropometric and adiposity-related traits, which are themselves risk factors for cardiovascular health; several causes of death in the UK Biobank (e.g. heart failure, vascular disease); and, unsurprisingly, blood protein, lipid and metabolite levels, as well as various red blood cell and immune cell counts.

As these results are association-based, they do not confirm the causal direction of the relationship between protein level and disease phenotype. Similarly, the observation that the lead variant at a pQTL is associated with another trait provides no evidence that the same variant is causal for both traits. An alternative approach to detect evidence of shared genetic risk variants, rather than these single-SNP associations, is to look systematically across the whole genome to see if alleles that increase plasma protein levels also increase disease risk.

### Genetic Correlations

To investigate if our proteins share genetic architecture with complex traits or cardiometabolic risk factors, we used High definition likelihood^33^ to estimate genetic correlations of our proteins with 14 important risk factors or outcomes (Supplementary Table 8, full results are in Supplementary Figure 7). Genetic correlations that remained statistically significant after Bonferroni correction for multiple testing (p<1.95×10^−5^) are shown in Figure 3. Interestingly, the traits with the most significant correlations with protein levels were BMI, WHR, creatinine and T2D; for BMI, WHR and T2D the majority were positive correlations, whereas all significant correlations between protein levels and creatinine were negative. IGFBP-2 levels are significantly genetically correlated with the most (8) traits including: lower BMI, WHR, total triglyceride levels, Type II diabetes and creatinine, but with increased HDL levels.

**Figure 3.**
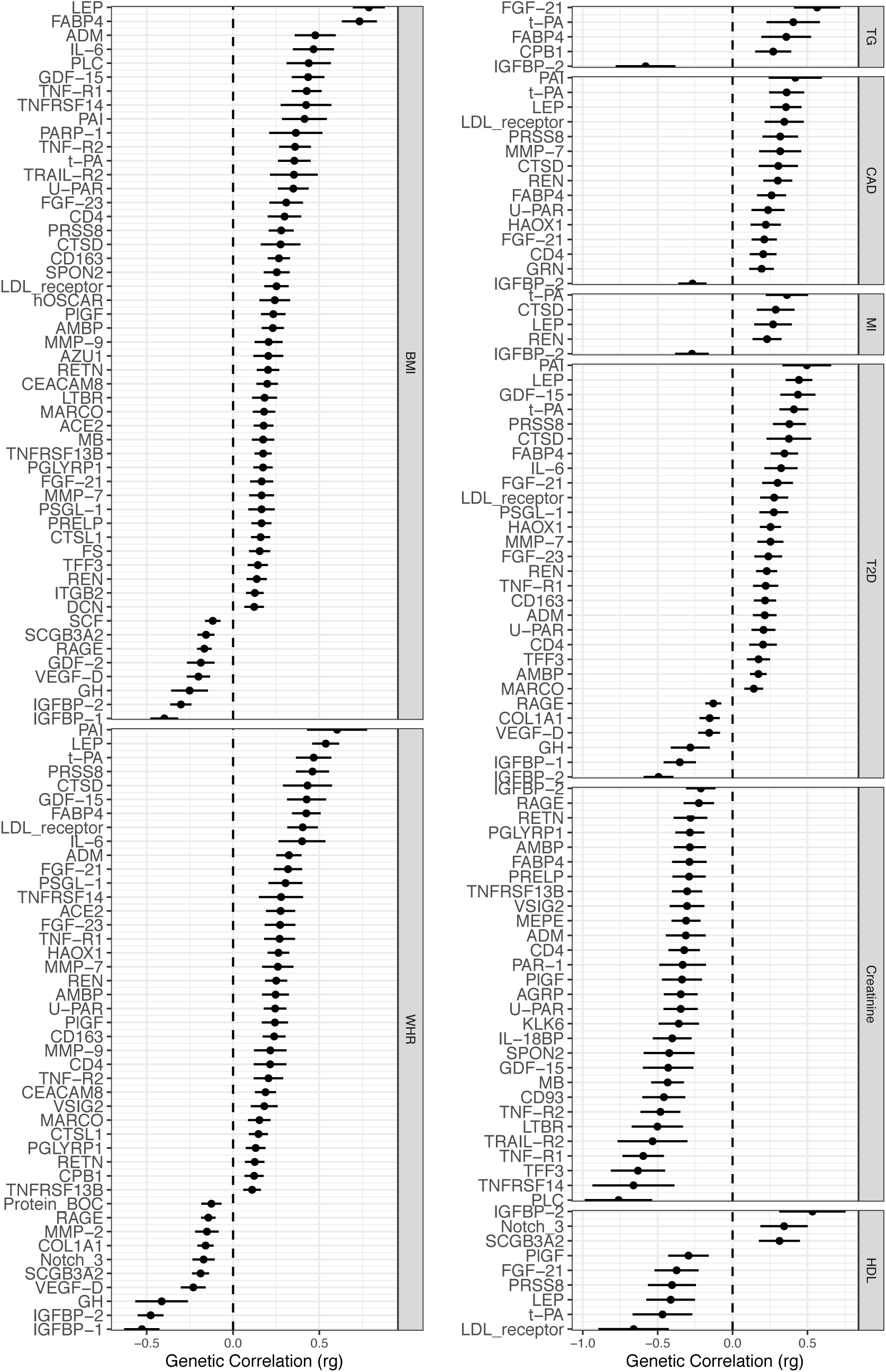
Genetic correlations show shared architecture between plasma protein levels and complex traits. Genetic correlation coefficients (rg) calculated using High definition likelihood for protein levels and complex traits. Only traits passing Bonferroni significance (p<1.95 × 10^−5^) are included (full results in Supplementary Figure 7). Error bars indicate 95% confidence intervals of rg estimation. BMI: Body mass index, WHR: waist-to-hip ratio, TG: triglyceride level, CAD: coronary artery disease, MI: myocardial infarction, T2D: type II diabetes mellitus, HDL: high density lipoprotein.

Several genetic correlations recapitulated known relationships. For example, we find that leptin levels (LEP) are genetically correlated with increased BMI, WHR, type II diabetes, coronary artery disease (CAD) and risk of myocardial infarction (MI), and lower HDL levels. This finding recapitulates known biology as leptin is involved in the regulation of energy homeostasis and is linked to type II diabetes and cardiovascular phenotypes^34^.

We also discovered novel correlations. The levels of LTBR (lymphotoxin beta receptor) circulating in the plasma were genetically correlated with increased BMI while BOC (Brother of CDO) and VSIG2 (V-set and immunoglobulin domain containing 2) levels correlated with WHR. None of these three proteins has been previously associated with adiposity-related traits.

In summary, our plasma protein levels share risk variants across the genome with health-related risk factors and disease outcomes, although an important caveat is that we are unable to distinguish the direction of these relationships from this analysis.

### Causal inference using Mendelian Randomisation

To identify potential causal relationships between plasma protein levels and disease we used Mendelian Randomisation (MR). We limited our analysis to only using *cis*-associated variants as instrumental variables (IVs) to reduce the influence of pleiotropy on our results. We also excluded any variants in the highly pleiotropic *HLA* and *ABO* regions. An LD threshold of r^2^>0.001 was used to remove correlated variants. We tested the association of the 169 proteins that had IVs meeting these criteria with 121 outcomes available from MR-Base^35^ (outcomes listed in Supplementary Table 9). 96 protein-outcome causal effect estimates passed a 1% FDR (Benjamini-Hochberg method^36^) significance threshold.

MR relies on assumptions that are difficult to test empirically. To increase confidence in our results, we therefore performed additional sensitivity analysis (Supplementary Table 10). To test the consistency of the causal estimates across IVs, we excluded any protein-outcome pairs if there was evidence of significant heterogeneity using Cochran’s Q test (q-value <0.05)^37^. To limit the chance of reverse causality influencing our results we performed bidirectional MR^38^ and excluded protein-outcome pairs that had significant causal effect estimates of outcome on protein level (p<3.62 × 10^−6^). For proteins with multiple *cis* IVs, we performed the pleiotropy-robust method MR-Egger^39^. An MR-Egger intercept estimate that is significantly different from zero can be interpreted as indicative of horizontal pleiotropy^39,40^. We therefore excluded protein-outcome MR estimates that had MR-Egger intercept p-values <0.05, leaving 59 significant protein-outcome causal estimates. Finally, to distinguish causal relationships from confounding due to LD we performed colocalisation analysis to look for evidence of a shared causal variant underpinning the genetic associations with protein and outcome. We used coloc^28^ and only considered protein-outcome pairs with a posterior probability (PP) of >0.8 of the hypothesis of a shared causal variant. We report 20 protein-outcome causal effect estimates that meet all of these criteria, involving 11 proteins associated with 16 different outcomes (Figure 4).

**Figure 4.**
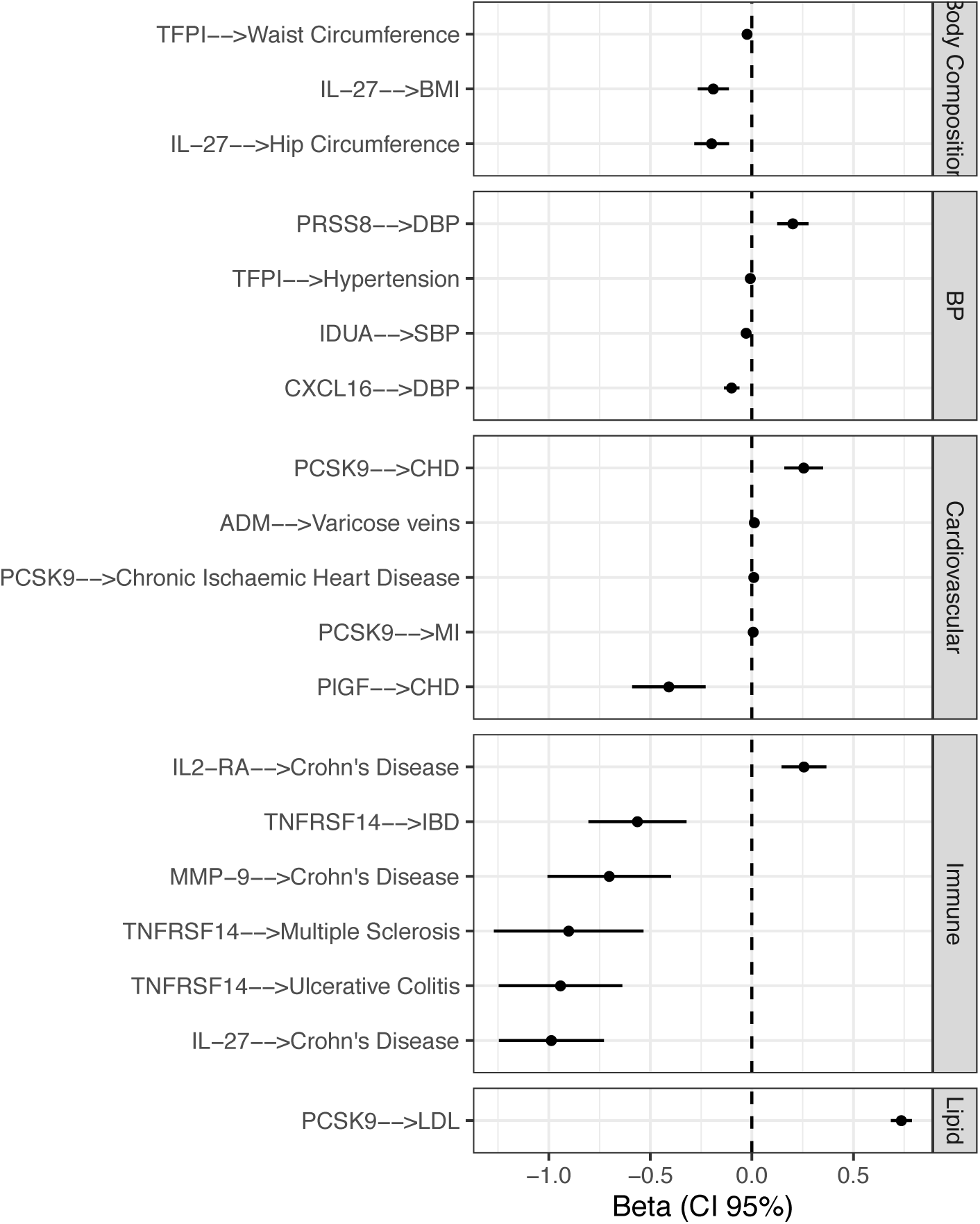
Cis-Instrument Mendelian Randomisation of plasma protein levels on complex diseases and health-related risk factors. MR causal effect estimates and 95% confidence intervals of the effect of plasma protein levels on outcome. Results from the fixed effects Inverse variance-weighted (IVW) method that passed 1% FDR, had a heterogeneity Q-value >0.05, an MR-Egger intercept p-value of >0.05, as well as evidence of a shared causal variant from colocalisation analysis (posterior probability of a shared causal variant >0.8) are shown. Associations are grouped by type of outcome. Causal effect estimates from additional MR methods that are robust to horizontal pleiotropy and relax the assumption of IVW allowing correlations between genetic associations with the exposure and outcome are in Supplementary Figure 8, as further sensitivity analyses. BMI: body mass index, DBP: diastolic blood pressure, SBP: systolic blood pressure, CHD: coronary heart disease, MI: myocardial infarction, IBD: inflammatory bowel disease, LDL: low density lipoprotein cholesterol levels.

The significant MR causal effect estimate of increased PCSK9 levels increasing LDL cholesterol levels (beta:0.74, SE:0.026) provides validation of the approach since the causal relationship of PCSK9 levels and LDL and total cholesterol levels is firmly established^41^; pharmacological inhibition of PSCK9 results in dramatic reductions in LDL cholesterol. In addition, our MR analysis confirmed previous reports indicating that PCSK9 increases risk of cardiovascular disease. This result is consistent with the findings of reduction in cardiovascular events in randomised clinical trials of PCSK9 inhibitors^42^. We also replicated other results from previous MR studies examining the role of circulating proteins in cardiovascular diseases and traits. This included the finding that a genetic tendency to higher placenta growth factor (PlGF) protein levels decreases the risk of CHD^18^, and that a genetic tendency to higher C-X-C Motif Chemokine Ligand 16 (CXCL16) protein levels decreases diastolic blood pressure^43^.

Variation in the genes encoding several of the proteins examined in our study have been associated with particular phenotypes. For example, SNPs mapped to Serine Protease 8 (*PRSS8)*, Interleukin 2 Receptor Subunit Alpha (*IL2RA)* and Tissue Factor Pathway Inhibitor (*TFPI)* have been associated with DBP^44^, Crohn’s disease (CD)^12,45,46^ and waist circumference^44^, respectively. Here, we advance these associations by demonstrating likely causal relationships between the circulating protein and the corresponding phenotypes through MR for the first time.

Our MR analysis provides novel insight into the pathogenesis of inflammatory bowel disease (IBD), which encompasses Crohn’s disease (CD) and ulcerative colitis (UC). IL27 is a heterodimeric cytokine that has complex biological functions including both pro- and anti-inflammatory effects in the intestine. IL27 can inhibit differentiation of Th17 cells, an important cell type in the pathogenesis of IBD. There are conflicting data on IL27’s role in IBD. In most^47,48^, although not all^49^, murine models of gut inflammation, IL27 is protective: IL27R genetic knockout worsens colitis while exogenous administration of IL27 ameliorates disease. In patients with IBD, *IL27* gene expression is elevated compared to controls^50^. Here we show that, in contrast to the observational human data, a genetic tendency to higher circulating IL27 is associated with lower risk of CD. This raises the possibility that the IL27 elevation in IBD patients results from reverse causation, perhaps as a response to dysregulated gut inflammation. Our data is in keeping with the observation that individuals with the risk allele for CD have lower IL27 gene expression^51^. Together this supports the concept that IL27 acts to protect the gut from aberrant inflammatory responses and raises the possibility that IL27 might be of therapeutic benefit in IBD.

By evaluating whether proteins play a causal role in disease aetiology, MR provides a valuable tool to identify and validate potential drug targets before embarking on costly clinical trials. We therefore examined whether any of the 11 proteins with inferred causal relationships in our MR analysis (Figure 4) were already current targets, using the DrugBank database^52^. In addition to PCSK9, which, as described previously, is a target of existing drugs used successfully in the treatment of hypercholesterolaemia and cardiovascular disease^42,53–55^, we found that 5 other proteins: PlGF, PRSS8, IL2-RA, MMP-9 (Matrix Metallopeptidase 9) and TFPI are also targets for drugs in various stages of development (Supplementary Table 11).

Our results highlighted IL2-RA as a potential candidate for drug repurposing. IL2-RA is the target for three approved drugs, two of these: Denileukin diftitox and Basiliximab, inhibit IL2-RA and are used for cutaneous T-cell Lymphoma (CTCL)^56^ and to prevent kidney transplant rejection^57^, respectively. The third, Aldesleukin, is an agonist and increases IL2-RA activity, inducing the adaptive immune response in the treatment of renal cell carcinoma^58,59^. Basiliximab has been piloted for use in IBD (UC) patients with apparent success in an uncontrolled open-label study^60^, however no benefit was found in an RCT^61^. Our finding that genetically increased levels of IL2-RA protein increase risk of CD (Beta: 0.26, SE: 0.06) suggest that further investigation is warranted into whether the suitability of Basiliximab (given the previous contradictory findings) may have a role in the management of CD.

Our inference that genetic predisposition to elevated MMP-9 decreases the risk of CD (Beta: -0.7, SE: 0.15) aligns with previous GWAS results: SNPs mapped to the *MMP9* gene have been associated (p<5 × 10^−8^) with lower risk of CD^12,46^. These genetics results are contrary to previous observational findings that increased serum MMP-9 levels were prognostic of clinical flare ups in CD patients^62^. Since MR is less prone to confounding and reverse causation than observational studies, we hypothesise that raised MMP-9 levels during flares of CD are likely to arise from reverse causation, perhaps reflecting an injury response. In keeping with this the MMP-9 inhibitor, Andecaliximab, was ineffective in phase 2 trials^63^, as would have been predicted by MR. This example highlights how integrating genetics and proteomics can be useful in deprioritising therapeutic targets.

We demonstrate the novel finding that MR identifies TNFRSF14 (HVEM) as protective against multiple immune-mediated diseases (IBD and MS). Notably, MS is also associated with polymorphisms in the TNFSF14 gene region, which encodes LIGHT, the ligand for TNFRSF14. The MS risk allele at TNFSF14 (LIGHT) is associated with lower serum levels of this protein^64^. This, together with our data, demonstrate that lower levels of both the ligand and its receptor are protective against MS, clearly indicating a causal role for this pathway in the maintenance of immune tolerance and raising the possibility that it could be manipulated for therapeutic benefit.

## Discussion

We have performed the largest pQTL study (Max N=26,494) on 184 plasma protein levels to date and report 592 independent loci significantly associated with the levels of at least one protein (1,308 protein-lead variant associations), with 687 lead variant-protein associations being novel. We found that estimates of the proportion of pQTL that overlap with eQTL ranges from 8.2-36.5% using multiple publicly available eQTL datasets and methods. Our results highlight that the majority of pQTL do not appear to be explained by eQTL. Given this finding, we highlight other potential mechanisms of action such as regulation of ligand-receptor pairs and transcription factor binding. The genetic architecture of plasma protein levels varies across proteins, from entirely polygenic (NEMO, GH) to single loci explaining almost all of the estimated genetic component (IL6-RA). Plasma protein levels also share genetic architecture with health-related risk factors and complex traits, with 52 protein levels being genetically correlated with BMI and 21 sharing heritability with CAD and MI. We also performed an extensive exploratory MR analysis using *cis*-pQTL as instruments, and found significant causal effect estimates for the levels of 11 proteins on 16 different outcomes. Our MR analysis highlighted plasma proteins that are candidate novel therapeutic targets and a candidate for drug repurposing.

In line with the larger size of our study, the discovery of a significant *cis*-pQTL for 92.7% of the plasma protein levels (where we tested the *cis*-regions) surpasses previous GWAS of plasma protein levels (18.5% Sun *et al*. N∼5,000, 86% Folkersen *et al*. N∼15,000)^4,18^. Additionally, CD93, ICAM-2, IL-6, PECAM-1 and t-PA levels had variants with p-values passing the genome-wide significance threshold for *cis*-signals (p<1×10^−5^) but were lost after correction for multiple testing, suggesting that our analyses were still underpowered and further *cis*-pQTL could be found in larger studies. Other than an issue of power, it is possible that our definition of *cis* (±1 Mb surrounding the coding region of the gene encoding the protein) is not capturing all signals however, no additional signals were found when the *cis*-region was widened to ±2 Mb. Ep-CAM, CD93, HAOX1, ICAM-2, MB, PECAM-1 and SRC proteins are intracellular^65^, which could contribute to significant signals not being found in samples from plasma. Expanding on previous studies, 78% of our significant pQTL were *trans*-associated compared to 68% in Folkersen *et al*. and 72% Sun *et al*. This is most likely due to our increased sample size, as like the aforementioned studies we found that proteins tended to have at most about 3 *cis*-pQTL, with any additional pQTL being *trans*-associated (Supplementary Figure 2), indicating that the increase in power allows the discovery of further *trans*-signals, which are likely to have smaller effect sizes (Welch T Test Two-sided p-value=1.48 × 10^−9^).

Akin to findings by Folkersen *et al*.^18^, we found that proteins varied in terms of their genetic architecture, with some proteins almost monogenic while others have polygenic architecture.

In terms of eQTL/pQTL overlap, our results based on direct lookup of lead variants found that 36.5% of our *cis*-pQTL had been previously reported as significant *cis*-eQTL (5% FDR). This is comparable to the 26% overlap based on variant lookup reported by Folkersen *et al*. and the 40% (including proxies LD r^2^≥0.8) by Sun *et al*. However, only 8.2% and 14.4% of our *cis*-pQTL showed strong evidence of colocalisation (PP>0.8) with the eQTL for the corresponding gene in eQTLgen and GTEx (at least one tissue), respectively. Additionally, coloc assumes that a single causal variant, included in the analysis, is driving the association signal in the region being considered. Given the strength of some of our *cis*-pQTL in particular, it is possible that the assumption of only one independent association signal could be violated. We limited the pQTL regions to ± 200 kb flanking the lead variant in our analysis, to minimise the chance of including multiple association signals. However, our findings are considerably lower than the reported 78.5% of 228 testable pQTL that showed evidence of colocalisation with eQTL in at least one tissue (PP>0.8) by Sun *et al*. One reason for this could be due to the difference in study design. Coloc assumes that the populations used to derive association statistics for the two traits have the same underlying pattern of LD. By meta-analysing multiple different populations, the LD structure in our sample will be different from those used to generate the eQTL datasets, whereas *Sun et al*. used only the INTERVAL cohort of English blood donors which may be a closer match to the GTEx population which was the source of their eQTL comparison. Recent methods that allow for multiple causal variants could overcome some of these issues. For example, the sum of single effects (SuSiE) regression framework coloc method^66^, however, this approach does require LD matrices for both populations and the use of a reference such as UK Biobank or 1000 Genomes will still not completely capture the LD structure in a multi-cohort GWAMA sample.

More generally, there are several reasons why colocalisation approaches might fail to indicate eQTL/pQTL overlap other than eQTL and pQTL having two independent causal variants: namely, differences in: sample size, assay technology or tissues between the two traits. The issue of tissue of origin is of particular concern here as, despite plasma having benefits as a medium, it does not accurately capture the levels of proteins in the tissues or cell types in which they are expressed and subsequently secreted, an inherent limitation when drawing conclusions about mechanisms. It is likely that higher eQTL/pQTL overlap would be observed if high-powered eQTL or pQTL datasets were available for the tissues from which the genes encoding these proteins are expressed. Despite GTEx having multiple different tissues, the small sample size means that its power is limited. This could also contribute to the low number of apparent tissue-specific overlapping eQTL/pQTL found using GTEx, as only those strong and robust *cis*-eQTL that are shared between tissues were able to be detected.

Our finding that 74.7% of our pQTL have been previously reported as DNA methylation QTL (meQTL) mirrors previous findings that 82% of *cis*-pQTL^17,20^ are also *cis*-meQTL^67^. These results highlight the link between DNA methylation and regulation of protein expression and exploring the interaction between plasma proteins and the epigenome would be an interesting avenue for further study, as would whether pQTL act by influencing mRNA splicing.

We restricted our MR analysis to *cis* IVs only, in contrast to previous studies^7,43,68^. This decision was made to fully take advantage of the direct biological link between *cis*-pQTL and protein level and to prevent highly pleiotropic *trans*-pQTL influencing our results by breaking the assumptions underlying MR. A systematic assessment of *cis* vs *trans* IVs would require the use of all of the most recent MR methods^69,70^ and meaningful results would be lost due to multiple testing. Additionally, we performed sensitivity analysis in line with the procedure set out by Zheng et al^43^, for using pQTL as IVs and showed that our causal effect estimates were consistent across multiple MR methods with varying assumptions (Supplementary Figure 8), therefore increasing confidence in the robustness of our results^68^.

Our exploratory MR analysis for plasma protein levels with a broad range of outcomes using the *cis* IVs was able to recapitulate the well-documented causal associations (PCSK9 with LDL and total cholesterol levels) and replicate findings reported by previous pQTL MR studies: genetically increased levels of CXCL16 and PlGF decreasing DBP^1^ and the risk of CHD respectively^2^. We also found evidence of novel causal associations between circulating protein levels (PRSS8, IL2-RA, TFPI and IL-27) and traits where the corresponding gene was already known to be associated, and novel causal protein-outcome relationships for ADM and IDUA. Using pQTL as IVs also highlighted IL2-RA as a potential candidate for drug repurposing and TNFRSF14 (IBD and MS) as novel therapeutic targets. Together these findings demonstrate the strength of our *cis*-pQTL as IVs and the potential for future discoveries by disease-specific analyses using this resource^9^.

Our increased sample size compared with previous pQTL studies^3,4,7,18^, is a particular strength of this work as it allowed us to discover novel pQTL for use as instruments. Additionally, the breadth of our approach exhibits the range of possible downstream uses of GWAS of circulating plasma protein levels. However, this breadth is also a limitation, as our work has uncovered numerous findings that inspire further research. For example, we found a significant proportion of pQTL did not overlap with the corresponding eQTL. This could be due in part to pQTL acting to influence the protein levels via other mechanisms such as influencing translation, clearing of the protein, export or expression of the protein’s receptor. However, this could also be due to the predominant use of whole blood eQTL datasets and the limited power of the multi-tissue dataset (GTEx), given that our proteins are also secreted in several tissues other than blood. Further analyses using high-powered eQTL datasets from the relevant tissues would be required to untangle the mechanisms of action of these pQTL. Similarly, we emphasise that the potential therapeutic targets identified by MR are preliminary and extensive investigations into other factors (e.g. druggability, safety) will also play a key role in determining the suitability of therapeutic intervention. As novel targets, TNFRSF14, ADM and IL-27, are either secreted into blood or retained membrane-bound or intracellular, dependent on isoform, further research into the specific functions of different isoforms is needed to validate their candidacy.

Our work builds on previous pQTL studies using a larger sample size for more proteins allowing the discovery of 1,308 significant protein-locus associations. By studying the genetic architecture of plasma protein levels, we have provided insight into the genetic regulation of protein levels, disease aetiology and casual relationships between circulating protein levels and cardiovascular disease phenotypes. In highlighting the power of our pQTL as IV to uncover candidate novel therapeutic targets in a broad exploratory analysis, we showcase the potential of this study as a resource to drive highly targeted research questions in the future.

## Data Availability

Meta-analyses summary statistics will be made publicly available upon publication. METAL software for meta-analysis is available from http://csg.sph.umich.edu/abecasis/metal/download/. SMR-HEIDI is available from https://cnsgenomics.com/software/smr/#Download. The Coloc R package is available from https://github.com/chr1swallace/coloc.

## Author Contributions

E.M.D contributed to the meta-analysis, visualisation of results. E.M.D, L.F, S.G, J.Z, N.E, A.R, S.E, N.M.C, D.V.Z, A.K., M.M, E.W, S.J.H, Y.C, A.P.M, B.P, U.V, N.J.W, J.D, J.S, B.G, D.B, R.J.S, H.C, U.G, C.Y, D.Z, T.L.A, P.E, D.L, C.L, J.G.S, T.E, J.F, O.H, Å.J, C.H, L.W, A.S, L.L, A.S.B, K.M, J.E.P and A.M, contributed to the cohort level analysis. E.M.D, L.K and P.R.H.J.T: contributed to other downstream analysis. E.M.D, P.K.J, J.F.W and J.E.P, contributed to manuscript drafting and editing. E.M.D, J.F.W and A.M contributed to project conception. P.K.J and J.F.W contributed to project supervision. All other authors commented and approved the manuscript prior to submission.

## Acknowledgements

The proteomic work was carried out under the aegis of the SCALLOP consortium. We thank Aikaterini Siopi for administrative assistance. We thank the UK Biobank resource, approved under application 19655. Cohort-specific acknowledgements can be found in Supplementary Table 12. J.F.W. and C.H, acknowledge support from the MRC Human Genetics Unit programme grant “Quantitative traits in health and disease” (U.MC_UU_00007/10). The work of L.K. was supported by an RCUK Innovation Fellowship from the National Productivity Investment Fund (MR/R026408/1). E.M.D. acknowledges the MRC Doctoral Training Programme in Precision Medicine (MR/N013166/1). RJS is supported by a UKRI Innovation-HDR-UK Fellowship (MR/S003061/1). We acknowledge the Swedish national research infrastructure SIMPLER (https://simpler4health.se/) for provisioning support in the analyses of COSM-C and SMCC. SIMPLER receives funding through the Swedish Research Council under the grant no 2017-00644.

## Disclaimer

The views expressed in this manuscript are those of the authors and do not necessarily represent the views of the National Heart, Lung, and Blood Institute; the National Institutes of Health; or the U.S. Department of Health and Human Services. The views expressed are those of the author(s) and not necessarily those of the NIHR or the Department of Health and Social Care.

## Data Availability

Meta-analyses summary statistics will be made publicly available upon publication.

## Code Availability

METAL software for meta-analysis is available from http://csg.sph.umich.edu/abecasis/metal/download/. SMR-HEIDI is available from https://cnsgenomics.com/software/smr/#Download. The Coloc R package is available from https://github.com/chr1swallace/coloc.

## Methods

### Proteomics Assay

Participating cohorts performed protein measurement using an antibody-based proximity extension assay (Olink Bioscience, Uppsala, Sweden)^75^ from EDTA plasma in 2 × 92-protein panels: ‘cvd2’ and ‘cvd3’. These targeted assays contained promising cardiovascular related proteins that also had two specific antibodies available for different epitopes. Analysis of all cohorts were conducted at one of two core laboratories with Olink Bioscience of SciLifeLab in Uppsala, Sweden.

### Statistical Analysis

#### Genome-wide Association

Summary statistics were obtained from 18 cohorts of European ancestry. Details of which cohorts contributed data for each protein are in Supplementary Table 13. The maximum sample size across all proteins was 26,494 however, average per-protein maximum and mean sample sizes were 23,981 and 18,141 respectively. It is worth noting that the CCL22 GWAS had a considerably smaller sample size (Max N=7460) than the other proteins as it was removed from the CVDIII panel by Olink during the data collection phase of this study, meaning only a subset of contributing cohorts returned CCL22 summary statistics.

The majority of cohorts provided data imputed with 1000 Genomes Project phase 3 or higher or to the Haplotype Reference Consortium (HRC) reference (Supplementary Table 12). Cohorts applied quality control filters for call rates, gender mismatch, cryptic relatedness and ancestry outliers. Cohorts performed genome-wide association studies on the inverse rank normalised NPX values. Below lower-limit-of-detection values (<LOD) were included in the analysis. Cohorts ran linear models adjusting for study-specific covariates such as batch or genotyping array as well as: age, sex, first 10 principal components of the genotypes to account for population structure, plate number, plate row, plate column, sample time in storage (days) and season of venepuncture. Studies containing related individuals corrected for kinship.

#### Meta-analysis

METAL^76^ software was used to perform inverse-variance-weighted meta-analysis (STDERR scheme) with the additional filters that only variants with an imputation quality score >0.4 and that were assessed in three or more cohorts were included. Heterogeneity of variant effect estimates between cohorts were also calculated using METAL.

#### Locus definition

In order to prevent heterogeneity influencing our results, only variants that had an I^2^<30% or have both: i) effect direction consistent with the meta in at least 3 individual cohorts and ii) be nominally significant (p<0.05) in at least 3 individual cohorts, were eligible to be considered genome-wide significant. Separate significance thresholds pre-correction for multiple testing were used for *cis*- (1 × 10^−5^) and *trans*-variants (5 × 10^−8^). A more liberal threshold was used for *cis-*signals as by only testing variants in the *cis*-region rather than genome-wide, fewer tests were performed. As the protein levels are correlated, rather than correcting the significance threshold for 184 traits, we calculated the number of PCs required to explain 95% of the variance in the 184 protein levels and took this value as the number of independent traits tested, as done previously by Kettunen *et al*.^77^. We found that 85 PCs explained 95% of the variance in the levels of 184 protein in ORCADES (using the “prcomp” function in R), we repeated the analysis in CROATIA-Vis and again found that 85 PCs explained 95% of the variance. Our thresholds for significance were therefore 1.18 x1 0^-7^ (Bonferroni 1 × 10^−5^/85) for *cis*- and 5.9 × 10^−10^ for *trans*-associated variants.

In order to identify non-overlapping loci associated with a given protein, 1 Mb windows were created around every significant variant for that protein. Starting with the region with the lowest p-value, any overlapping windows were then merged, this was repeated until no more 1 Mb windows remained. To refine a list of non-overlapping loci that are associated with at least one of our 184 proteins we repeated this process of merging overlapping 1 Mb windows on the list of significant protein-locus associations.

#### Conditional Analysis

Conditional analysis was performed per protein using the --cojo-slct method from GCTA-cojo^78^. A minor allele frequency (MAF) filter of 1% and a p-value threshold of 1 × 10^−5^ were used. A random 10,000 unrelated genetically genomically British individuals from the UK Biobank were used as linkage disequilibrium (LD) reference.

Due to the particularly strong *cis*-signals we further filtered conditional variants, retaining per protein those with r^2^<0.001. The criteria to limit heterogeneity for our primary variants were applied to conditionally associated variants, retaining those with I^2^<30 or if I^2^>30 then at least 3 cohort level results that have consistent effect direction with the meta-analysis and nominally significant at the cohort level (p<0.05). As with primary associated variants the threshold for significance corrected for multiple testing was 5 × 10^−8^/85 for *trans* variants and 1 × 10^−5^/85 for *cis* variants. Finally, akin to the primary variants, for each protein 1 Mb windows were created around each significant conditionally independent variant, with overlapping windows being merged, starting with the lowest p-value, until none are remaining.

#### Novelty of pQTL

To establish novelty of pQTL, we tested whether our 1,308 lead variants (or variants in LD, r^2^>0.5, with our lead variants) had been previously associated with the relevant protein in 22 published GWAS or plasma protein levels (Supplementary Table 4).

#### Heritability

Estimates of total SNP heritability for each circulating plasma protein level were calculated as the sum of the contributions from two independent partitions of the SNPs: pQTL and the polygenic component. The pQTL component was calculated as the sum of the estimated variance explained (VE) in protein level by the lead variants of the primary pQTL. VE for each lead variant was estimated as 2*pqβ*^*’*^where *β* is the meta-analysis effect size, *p* is the effect allele frequency and *q* = 1 − *p*. The polygenic component was estimated using linkage disequilibrium-score regression (LDSC)^22^ using variants present in the European 1000

Genomes Phase 3 Reference sample^79^. To ensure that variance explained by SNPs in LD with lead variants was not counted twice, variants within ±10 Mb of lead variants were excluded from calculations of the polygenic component.

#### Annotation of Significant Loci

Previously reported associations of all 1,093 of our significant variants and their proxies with an r^2^>0.8 based on a 1000 Genomes Phase 3 European reference with GWAS traits, eQTL, proteins, metabolites and methylation QTLs were extracted from Phenoscanner v2^32,80^, with a p-value threshold of 5 × 10^−8^. Lead variants were also queried for evidence of being in a regulatory region using RegulomeDB^29^.

For each of the *trans*-associated variants we defined a set of *trans* genes. These *trans* genes were any genes whose coding regions overlapped with a ± 500 kb window surrounding our significant variant using the Homo.sapiens^81^ annotation package in R. For each of the *trans* genes we looked to see if the protein they encode have any known interactions with the protein we found it associated with using the STRINGdb^30^ R package (database version 10). Similarly, for each *trans* gene we looked to see if they had any known pathways, gene ontology (GO) terms or publications in common with the gene encoding the protein we found them associated with. This was done using the KEGGREST^31^ and org.Hs.eg.db^82^ R packages.

#### Colocalisation of pQTL and eQTL

We looked up whether any of our significant variants had been previously reported as a significant eQTL (5% FDR (permutation-based)) in whole blood expression data from eQTLgen^24^ and from 48 different tissues using the Genotype-Tissue Expression project (GTEx) v7.

SMR-HEIDI^25^ was used to test whether a single causal variant is influencing gene expression and protein level due to either causality or pleiotropy, however it cannot distinguish between the two. We tested if ±500 kb regions flanking all 1,308 of our significant lead variants were associated with gene expression using four publicly available eQTL datasets: 48 GTEx tissues, both *cis* and *trans* eQTLs from eQTLgen^24^, Westra *et al*.^26^ and Cage^27^. Correction for multiple testing was carried out per eQTL dataset, with results with PSMR passing Bonferroni correction for number of proteins vs probes and PHEIDI≥0.01 considered significant.

In order to distinguish between causality and pleiotropy, we performed colocalisation using the “coloc.abf” function from the “coloc”^28^ R package, with default priors. This approach simultaneously calculates posterior probabilities (PP) of eQTL and pQTL i) sharing a single causal variant and ii) being driven by two independent variants. For each of our *cis*-pQTL, the region within ±200 kb of the lead variant was tested for colocalisation with the gene encoding the protein in both the eQTLgen and 48 tissues from GTEx v7. We considered a PP>0.8 for the hypothesis that eQTL and pQTL share a causal variant as strong evidence of colocalisation and a PP>0.5 as likely to colocalise^4^.

#### Genetic Correlations

The High definition likelihood^33^ R package was used to estimate genetic correlations between the levels of our 184 proteins and the following cardiovascular-related traits using publicly available summary statistics (Download URLs in Supplementary Table 8): body mass index (BMI), coronary artery disease (CAD), chronic obstructive pulmonary disease (COPD), creatinine levels, Crohn’s Disease, high density lipoprotein cholesterol (HDL), low density lipoprotein cholesterol (LDL), myocardial infarction (MI), Rheumatoid arthritis (RA), type II diabetes (T2D), total cholesterol, triglyceride levels and waist-hip ratio (WHR). To aid visualisation, proteins and complex traits were ordered using Euclidean distance-based hierarchical clustering with the hclust function in R.

#### Mendelian Randomisation

##### Instrument selection

For each protein, instruments were selected from genome-wide significant variants that passed the additional criteria of i) having a meta-analysis heterogeneity I^2^<30 or if I^2^>30, then ii) must have effect direction consistent with the meta-analysis in at least 3 cohorts and iii) be nominally significant (p<0.05) in at least three cohorts. These variants were then clumped for LD using an r^2^ filter of 0.001 with the “TwoSampleMR”^35^ R package. For each protein, MR was run using *cis* variants, with any variants within the HLA (chr6:29645000-6:33365000, build 37) and ABO (chr9:136131052-9:136150605, build 37) regions excluded from selection as instruments.

##### Primary MR Analysis

“TwoSampleMR” was used to perform Mendelian randomisation (MR) analysis. Protein level exposures were tested against 121 outcomes available in the MR-Base database^35^ (the full list of outcomes tested is in Supplementary Table 9) using the fixed effects inverse variance-weighted (IVW) method. Outcomes were selected due to their relation to cardiovascular disease risk or immune-related disorders, given the proportion of immune system-related proteins in our set. For each outcome, summary statistics with the largest sample size and closest ancestry match with our GWAMA population were chosen.

##### Sensitivity analyses

To minimise the risk of heterogeneity between IVs influencing our results, only those without evidence of significant heterogeneity, using Cochran’s Q test (q-value>0.05)^37^, were considered. Additionally, to limit the effect of horizontal pleiotropy, we excluded protein-outcome MR estimates that had MR-Egger intercept significantly deviating from zero (P<0.05)^39,40^. We also performed MR analysis using MR-Egger, weighted median and weighted mode methods, which are more robust to horizontal pleiotropy^69,83^ (Supplementary Figure 8). We also used the maximum likelihood (ML) method^84^ which relaxes the assumption used by the IVW method, allowing both: uncertainty in the effect size of the IVs with the exposure and correlations between the genetic associations with the exposure and outcome. Consistency in causal estimates across MR methods with varying assumptions increases the chance of robust results.

##### Colocalisation

To distinguish causal relationships from confounding due to LD, we tested for evidence of a shared causal variant between each protein-exposure outcome pair using colocalisation. Variants within ±200 kb of each IV were tested for colocalisation with the overlapping variants in the outcome GWAS (extracted from MR-Base using the “associations” function from the ieugwasr R package). Only those with a posterior probability estimate of >0.8 for hypothesis 4 were considered further. Sample sizes for the 26 outcomes in the 59 protein-outcome associations passing 1% FDR heterogeneity and pleiotropy filters ranged from N=462,116 to N=173,082, for quantitative traits (N=119,731 to N=7,735 cases, for binary traits).

##### Bi-directional analyses

We tested for evidence of causal associations of the 121 outcomes on proteins using the IVW method. Protein-outcome pairs that had a causal effect estimate with p<3.62 × 10^−6^ (Bonferroni 0.05/13,810) were not considered further due to the potential for the estimate for the effect of protein on outcome to be influenced by reverse causality.

#### Drug Targets

The DrugBank Release Version 5.1.7^52^ was used to see if the 11 proteins that had evidence of significant causal associations (PMR passed 1% FDR & additional criteria described above) causal associations in our MR analysis are current drug targets.

